# Forecasting COVID-19 and Analyzing the Effect of Government Interventions

**DOI:** 10.1101/2020.06.23.20138693

**Authors:** Michael Lingzhi Li, Hamza Tazi Bouardi, Omar Skali Lami, Thomas A. Trikalinos, Nikolaos K. Trichakis, Dimitris Bertsimas

**Affiliations:** Operations Research Center, Massachusetts Institute of Technology, Cambridge, MA 02139; Center for Evidence Synthesis in Health, Brown University, Providence, RI 02912; Sloan School of Management, Massachusetts Institute of Technology, Cambridge, MA 02142

**Keywords:** Epidemiology, COVID-19, Government Intervention, Underdetection, Reopening

## Abstract

One key question in the ongoing COVID-19 pandemic is understanding the impact of government interventions, and when society can return to normal. To this end, we develop DELPHI, a novel epidemiological model that captures the effect of under-detection and government intervention. We applied DELPHI across 167 geographical areas since early April, and recorded 6% and 11% two-week out-of-sample Median Absolute Percentage Error on cases and deaths respectively. Furthermore, DELPHI successfully predicted the large-scale epidemics in many areas months before, including US, UK and Russia. Using our flexible formulation of government intervention in DELPHI, we are able to understand how government interventions impacted the pandemic’s spread. In particular, DELPHI predicts that in absence of any interventions, over 14 million individuals would have perished by May 17th, while 280,000 current deaths could have been avoided if interventions around the world started one week earlier. Furthermore, we find mass gathering restrictions and school closings on average reduced infection rates the most, at 29.9 ± 6.9% and 17.3 ± 6.7%, respectively. The most stringent policy, stay-at-home, on average reduced the infection rate by 74.4 ± 3.7% from baseline across countries that implemented it. We also illustrate how DELPHI can be extended to provide insights on reopening societies under different policies.

## 1. Introduction

Currently, the world is facing the deadliest pandemic in recent history - COVID-19. As of December 16th, there have been over 74.2 million confirmed cases of COVID-19 and the disease has taken over 1,648,000 lives. To stop the further spread of COVID-19, governments around the world have enacted some of the most wide-ranging non-pharmaceutical interventions in history. These interventions, especially the more severe ones, carry significant economic and humanitarian cost. Thus, it is critical to understand the effectiveness of such interventions in limiting disease spread.

However, there are many challenges in attempting to understand the effect of government interventions in a specific region or country. Different regions have implemented, often concurrently, a variety of different policies, and worse, even the same interventions could produce largely different effects in different societies, due to differences in factors such as demographics, population density, and culture.

Thus, in order to provide a sensible analysis of the effect of policies across different countries, in late March, 2020 we created a novel epidemiological model, DELPHI, to model the spread of COVID-19. DELPHI (Differential Equations Lead to Predictions of Hospitalizations and Infections) extends a classical SEIR model (Kermack and McKendrick 1927) to include many realistic effects that are critical in this pandemic, including deaths, underdetection, and changing governmental interventions. Since its inception in late March, DELPHI has been one of the top 4 models consistently incorporated into the US Centers for Disease Control and Prevention’s (CDC) core ensemble forecast (Dean et al. 2020) and have been utilized by various health and federal agencies including the Federal Reserve for pandemic planning. A major hospital system in the United States, Hartford Healthcare, planned its intensive care unit (ICU) capacity based on our forecasts.

One of the key innovations of DELPHI is the explicit and parametric characterization of government interventions. This allows us to understand the effect of different non-pharmaceutical interventions as they have been implemented in various regions while accounting for regional population characteristics including baseline infection rate and mortality percentage. In Section 4.1, we provide evidence that school closings and mass gathering restrictions were among the most effective measures in reducing the rate of infection during the early stages of the pandemic, though they carry a significant social burden. This analysis suggests that despite their extreme cost, these policies were effective in controlling the extreme growth in infections when other preventative measures (e.g. masks) and treatment options were still being evaluated and developed. As a further illustration of their outsized effect, we are able to demonstrate that had these restrictions been implemented just one week earlier, most – up to 90%– of the deaths in the early stages of the pandemic could have been avoided.

Another major use case of DELPHI is scenario analysis for the future to enable long-term planning. In Section 5, we demonstrate how DELPHI were utilized by Janssen Pharmaceuticals (a Johnson & Johnson company) in mid-to-late 2020 to determine the Phase III trial locations of their leading vaccine candidate Ad26.Cov.S to maximize baseline incidence based on differing scenarios of governmental interventions.

DELPHI has been applied to 167 geographic areas (countries/provinces/states) worldwide as of end of April, and more than 215 as of end of September, covering all 6 populated continents. Its results and insights have also been available since early April on www.covidanalytics.io. In this paper, we document the statistical innovations, quantitative results, and insights extracted from the DELPHI model.

### 1.1. Literature

As the COVID-19 pandemic worsened, there has been a large number of research groups that created COVID-19 epidemiological models in an effort to help understand the crisis. A large class of models utilize some flavor of the SEIR compartmental model (Kermack and McKendrick 1927), which attempt to model large population dynamics by assuming the population divides into several compartments that have dynamics coupled through differential equations. Some of these models utilize a classical compartmental model (**?**) while others create subdivisions based on age groups and symptom severity to account for heterogeneity (DRA 2020). Furthermore, many modeling approaches supplement SEIR models with additional behavioral data such as mobility and governmental policies (Woody et al. 2020, Chinazzi et al. 2020) to adjust for compliance and non-pharmaceutical interventions. There is also significant work on approaches that do not model the disease dynamics directly, such as utilizing deep learning for predicting week-ahead mortality (Rodriguez et al. 2020), or treating the past epidemiological data as a time-series forecasting problem (Mehrotra and Ivan 2020). For a more comprehensive review of the different models that have been developed, we refer the reader to Dean et al. (2020).

The DELPHI model is based on the SEIR compartmental model but differs significantly with the previously illustrated models on multiple fronts. First, instead of utilizing governmental policies directly, we utilize an embedded parametric curve in the epidemiological model to model the effect of governmental policy. During this pandemic, we often observe that there is a large discrepancy between the official policy and the observed data on the ground, and thus this approach would allow us to capture the *experienced* effect of the policy as reflected by the epidemiological data. Similarly, parametric curves are utilized to model changes in mortality percentage due to advances in treatments and care for COVID-19. We also utilize an explicit separation of modeling for deaths and recoveries that allows us to fit to multiple end-points (recorded cases and deaths) simultaneously, in contrast to many approaches which build separate models for each endpoint (e.g. IHME 19).

Overall, the flexibility of the parametric curves allows us to be one of the very few models that are able to produce consistent projections on all 6 populated continents. Our parametric modeling of the infection rate also allows us to give actionable policy guidance by analyzing which non-governmental interventions were more effective as described in Section 4.1 and showcasing the degree of which early intervention could prevent the epidemic in Section 4.2.

Finally, the DELPHI model is one of the top 4 models to be consistently included in the CDC ensemble forecast (Dean et al. 2020), and its favorable performance is demonstrated in Section 3.2.

## 2. The DELPHI Model

The DELPHI model is a compartment epidemiological model that extends the classical SEIR model into 11 states under the following 8 groups:

- **Susceptible (*S*)**: People who have not been infected.
- **Exposed (*E*)**: People currently infected, but not contagious and within the incubation period.
- **Infected (*I*)**: People currently infected and contagious.
- **Undetected (*U***_***R***_**) & (*U***_***D***_): People infected and self-quarantined due to the effects of the disease, but not confirmed due to lack of testing. Some of these people recover (*U*_*R*_) and some die (*U*_*D*_).
- **Detected, Hospitalized (*DH***_***R***_**) & (*DH***_***D***_): People who are infected, confirmed, and hospitalized. Some of these people recover (*DH*_*R*_) and some die (*DH*_*D*_).
- **Detected, Quarantine (*DQ***_***R***_**) & (*DQ***_***D***_): People who are infected, confirmed, and home-quarantined rather than hospitalized. Some of these people recover (*DQ*_*R*_) and some die (*DQ*_*D*_).
- **Recovered (*R*)**: People who have recovered from the disease (and assumed to be immune).
- **Deceased (*D*)**: People who have died from the disease.

In addition to main functional states, we introduce auxiliary states to calculate a few useful quantities: Total Hospitalized (TH), Total Detected deaths (DD) and Total Detected Cases (DT). The full mathematical formulation of the model is as followed:

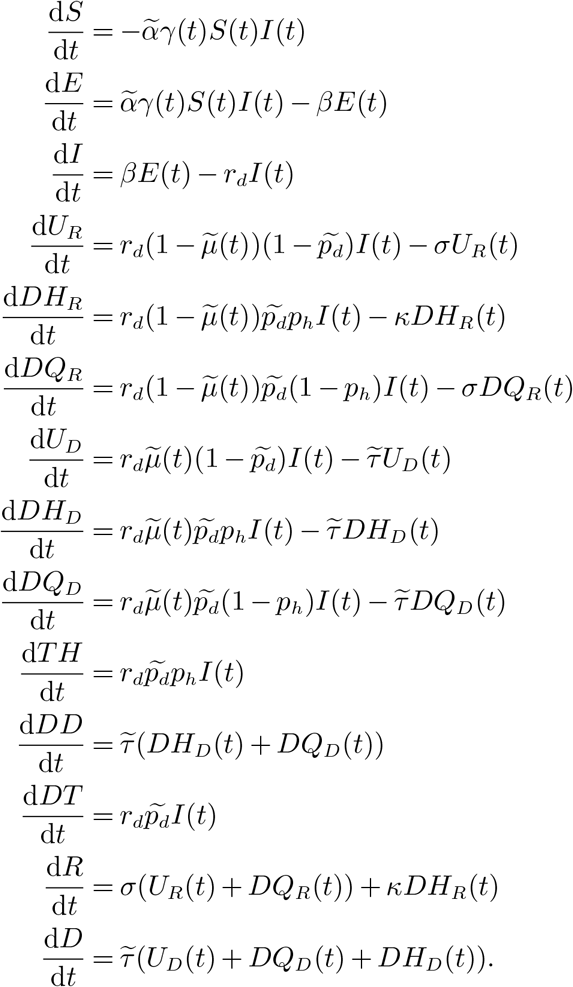

Figure 1a depicts a flow representation of the model, where each arrow represents how individuals can flow between different states. The underlying differential equations are governed by 11 explicit parameters which are shown on the appropriate arrows in Figure 1a and defined below. As the pandemic progressed, the DELPHI model was also continuously updated to reflect the changing situation. Definitions that have been updated during the course of the pandemic are detailed for full transparency. To limit the amount of data needed to train this model, only the parameters denoted with a tilde are being fitted against historical data for each area (country/state/province); the others are largely biological parameters that are fixed using available clinical data from a meta-analysis of over 190 papers on COVID-19 available at time of model creation (Bertsimas et al. 2020). A small selection of references for each parameter is given below.

**Figure 1.**
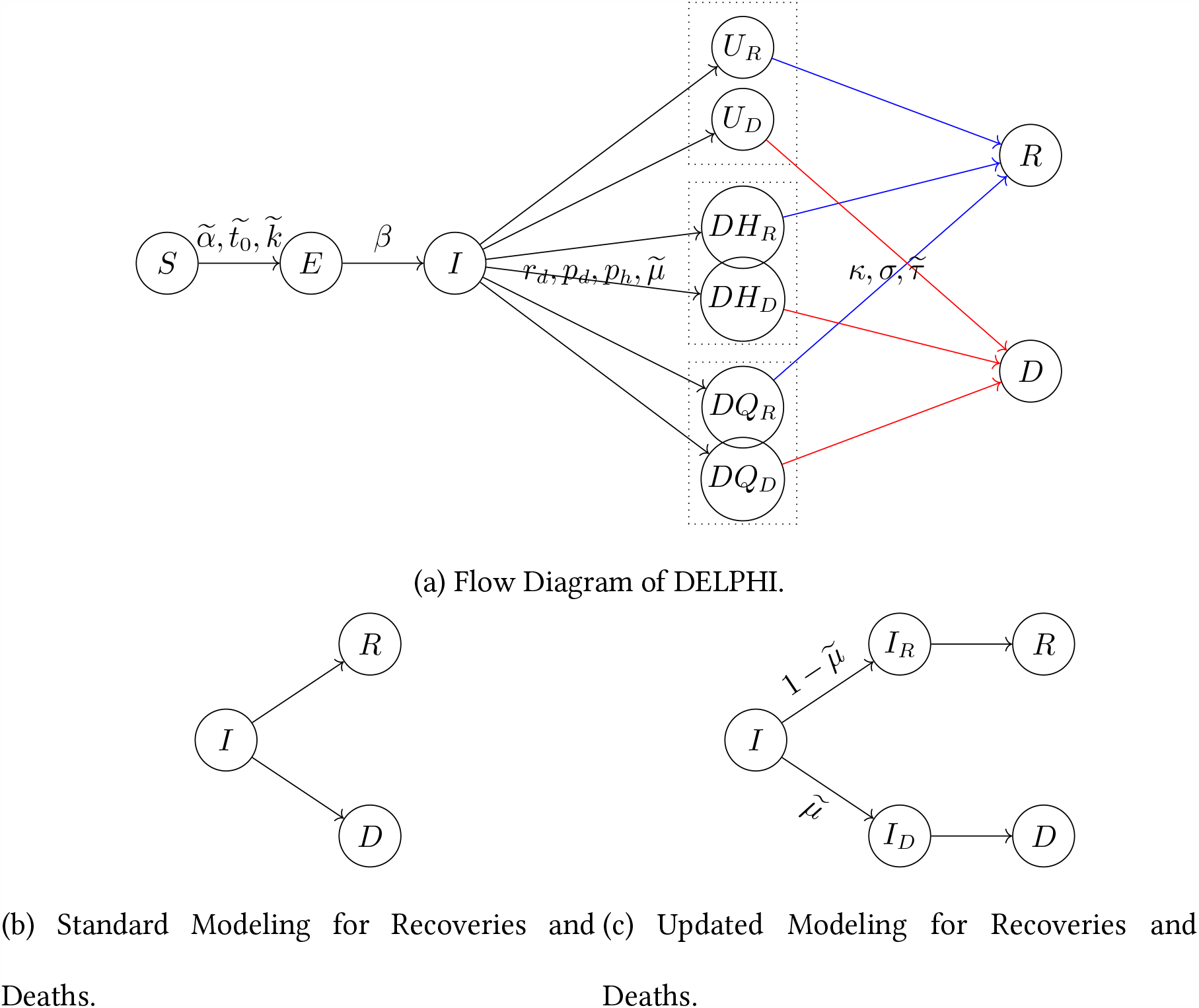
The DELPHI Model.

- 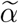 is the baseline infection rate.
- *γ*(*t*) measures the effect of government response and is defined as:

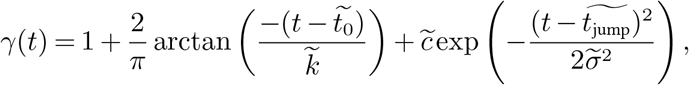

where the parameters 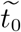 and 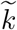 capture, respectively, the timing and the strength of the response. The exponential term intends to reflect a resurgence in infections due to relaxation of governmental policy and societal response, where 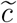 controls the magnitude of resurgence, 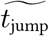 the time of the acme of the resurgence, and 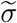 the duration of the resurgence phase. The effective infection rate in the model is 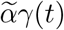, which is time dependent. The exponential resurgence term was added to the model in late June as we observed the large resurgence in the pandemic (*i*.*e*. before July the model assumed 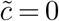). The 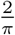 constant is so that the starting *γ*(*t*) with 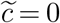 is normalized to the range of [0, 2] with *γ*(*t*) = 1 if *t* = *t*_0_.

- *r*_*d*_ is the rate of detection. This equals to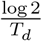, where *T*_*d*_ is the median time to detection (fixed to be 2 days), see Wang et al. (2020).
- *β* is the rate of infection leaving incubation phase. This equals to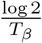, where *T*_*β*_ is the median time to leave incubation (fixed at 5 days), see Lauer et al. (2020).
- *σ* is the rate of recovery of non-hospitalized patients. This equals to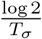, where *T*_*σ*_ is the median time to recovery of non-hospitalized patients (fixed at 10 days), see Hu et al. (2020), Kluytmans et al. (2020).
- *κ* is the rate of recovery under hospitalization. This equals to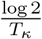, where *T*_*κ*_ is the median time to recovery under hospitalization (fixed at 15 days), see Liu et al. (2020b), Grein et al. (2020).
- 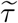 is the rate of death. This captures the speed at which a dying patient dies, and thus inversely proportional to how long a dying patient stays alive.
- 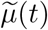 is the mortality percentage, defined as:

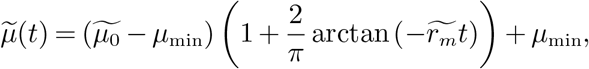

Where 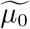 is the initial mortality percentage, *µ*_min_ is the minimum mortality percentage and 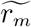 is the decay rate for mortality. Before June, the model had assumed 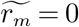 as the mortality percentage was relatively constant in the early pandemic due to the lack of treatment options. This parametric curve describes the natural decay of mortality percentage as standard of care improves throughout the pandemic. Notice that this quantity measures the percentage of people who die from the disease in a particular region, and is independent from the rate of death.

- *p*_*d*_ is the percentage of infectious cases detected, which is fixed at 20% based on various reports trying to understand the extene of underdetection in countries with earlier outbreaks. Wang et al. (2020), Krantz and Rao (2020), Niehus et al. (2020)
- *p*_*h*_ is the (constant) percentage of detected cases hospitalized, which is set to 15%, see Arons et al. (2020), Xu et al. (2020).

We fit on 11 parameters from the list above 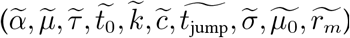. In addition, we introduce 2 additional parameters 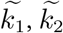 to account for the unknown initial population in the infected (*I*) and exposed (*E*) states. We thus fit 12 parameters per area.

The parameters are fitted by minimizing a weighted Mean Squared Error (MSE) metric with respect to the parameters. Let *DT* (*t*) and *DD*(*t*) denote the number of reported total detected cases and detected deaths, respectively, on day *t*. Then, the loss function for a training period of *T* days is defined as:

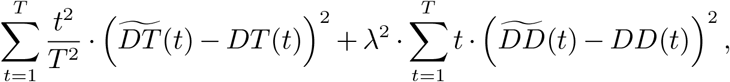

where 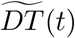 and 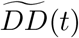 are respectively the total detected cases and deaths predicted by DELPHI. The factor 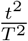 gives more prominence to more recent data, as recent errors are more likely to propagate into future errors. The lambda factor 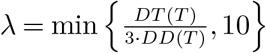 balances the fitting between detected cases and deaths; this re-scaling coefficient was obtained experimentally. We utilize a rolling training window of up to 4 months of historical data to train the model, as the parametric form of *γ*(*t*) would allow both the modeling for the current wave and any resurgence in the future. We specifically exclude historical data starting before the area recorded more than 100 cases; this allows us to exclude sporadic outbreaks that are not epidemics. To optimize over the highly non-convex search space, we utilize both the local truncated newton algorithm (Nocedal and Wright 2006) and the global optimization method of dual annealing (Xiang et al. 1997). The truncated newton algorithm is utilized to produce forecasts on a daily basis while the dual annealing optimization is performed on a weekly basis to shift and re-adjust the parameters more significantly if the underlying mechanics have changed (e.g. in the case of a new wave of cases). This dual-track optimization allows efficient optimization while ensuring that the resulting fit would not drift significantly from the data. We next discuss the three key characteristics of the DELPHI model compared to the standard SEIR formulation as well as the variants used for COVID-19 modeling which we have referenced in section 1.1.

### Accounting for Under-detection

In the COVID-19 crisis, one of the key modeling difficulties is the chronic underdetection of confirmed cases. This is both due to the lack of detection abilities in the early stages of the pandemic and also the similarity between a mild case of COVID-19 and the common flu. Thus, to account for such a significant effect, we explicitly included the *U*_*R*_/*U*_*D*_ states to model people who actually contracted COVID-19 (and are infectious), but were not detected. In particular, we assume that only *p*_*d*_ of the total number of the cases were detected, while 1 − *p*_*d*_ of the total cases flow to the *U*_*R*_/*U*_*D*_ states. There are two methods to gain information on the detection rate: treating *p*_*d*_ as a parameter and fit to the historical data, or recover *p*_*d*_ from serological evidence. However, both methods were impractical during the creation of this model. In an early to mid stage pandemic, a wide range of detection percentages are consistent with the data but leads to vastly different predictions (see e.g. Lourenço et al. (2020)), so historical data could not provide strong evidence. Furthermore, at the time of redaction, the serological data were largely limited to specific sub-areas such as cities and counties (see Doi et al. (2020), Streeck et al. (2020), Sood et al. (2020), Bendavid et al. (2020) for examples), while region-wide surveys were largely limited to a few European countries (see Erikstrup et al. (2020), Wise (2020) for examples and discussion) and only very sparsely available around the world. Note that testing data was not explicitly used in the differential equations of the DELPHI model, because trying to regress the coefficient *γ*(*t*) or the detection percentage on the time-varying tested percentage of the population significantly decreased the empirical performances of the model. This can be explained by the inconsistency of the reported testing across different regions, and the fact that tests were not uniformly distributed across the population as they were highly skewed towards the symptomatic individuals, especially in the earlier stages of the pandemic. One possible direction for improvement and future research is to smooth this testing data, for example using 7-day rolling averages, before feeding it to the DELPHI model.

Thus, we instead fix the detection percentage to be 20% based on various reports trying to understand the extent of underdetection in countries with earlier outbreaks (Wang et al. 2020, Krantz and Rao 2020, Niehus et al. 2020). Even more recently, Breton (2020) has independently reached a similar conclusion to our assumption. We acknowledge that this is a major simplification of the true dynamics as we expect the underlying detection rate to be changing over time and also in different areas; however, the sensitivity analysis conducted in Section 3.3 demonstrates that the model is relatively insensitive to this underlying parameter, which is unidentifiable.

### Separation of Recovery and Deaths

A large focus in many governments’ response to the COVID-19 pandemic is to minimize the number of deaths, and thus in DELPHI, we included a death state. In most epidemiological models that extend to include the death state (see e.g. Wang et al. (2020), Peng et al. (2020) for COVID-19 modeling examples), the death state (*D*) is shown to flow from the same active infectious state as the recovery state (*R*), with a schematic shown in Figure 1b. However, this modeling approach would cause the mortality percentage 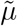 to be dependent on the rates of recovery and death.

Let us assume that recoveries and deaths are modeled as shown in Figure 1b. Then, the outflow from the *I* state, denoted *I* ^−^, can be written as

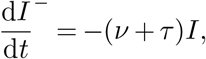

where *ν* is the rate of recovery and *τ* is the rate of death. This state dependent model implies a mortality percentage of 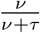, which is fixed given the rate of recovery and the rate of death. However, in reality, the mortality percentage should be independent from how fast people recover or die. Thus, to resolve such mismatch, we explicitly separated out the 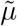 fraction of the population infected that would eventually die (*I*_*D*_) from the 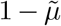 fraction that would recover (*I*_*R*_), as illustrated in Figure 1c.

This allows the mortality percentage 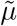 to be independent from the rates of death and recovery. The final DELPHI model further differentiated the *I*_*R*_ states into hospitalized (*DH*_*R*_), quarantined (*DQ*_*R*_), and undetected (*U*_*R*_) states to account for the different treatments people received, and similarly with the *I*_*D*_ states.

### Modeling Effect of Increasing Government Response

One of the key assumptions in the standard SEIR model is that the rate of infection *α* is constant throughout the epidemic. However, in real epidemics such as the COVID-19 crisis, the rate of infection starts decreasing as governments respond to the spread of epidemic, and induce behavior changes in societies. To account for such effect, we model the effect of government measures with a parametric function *γ*(*t*) that includes an arctan function, as well as an exponential function.

In total the parametric curve models 4 separate phases in a pandemic. The concave-convex nature of an arctan curve accounts for the first three phases: The early, concave part of the arctan models limited changes in behavior in response to early information, while most people continue business-as-usual activities. The transition from the concave to the convex part of the curve quantifies the sharp decline in infection rate as policies go into full force and society experiences a shock event. The latter convex part of the curve models a flattening out of the response as the government measures reach saturation, representing the diminishing marginal returns in the decline of infection rate. Then the exponential term is meant to model a potential resurgence in cases, for instance due to premature relaxation of societal restrictions or some other behavior in the area’s society. An illustration of such four phases is shown in Figure 2.

**Figure 2.**
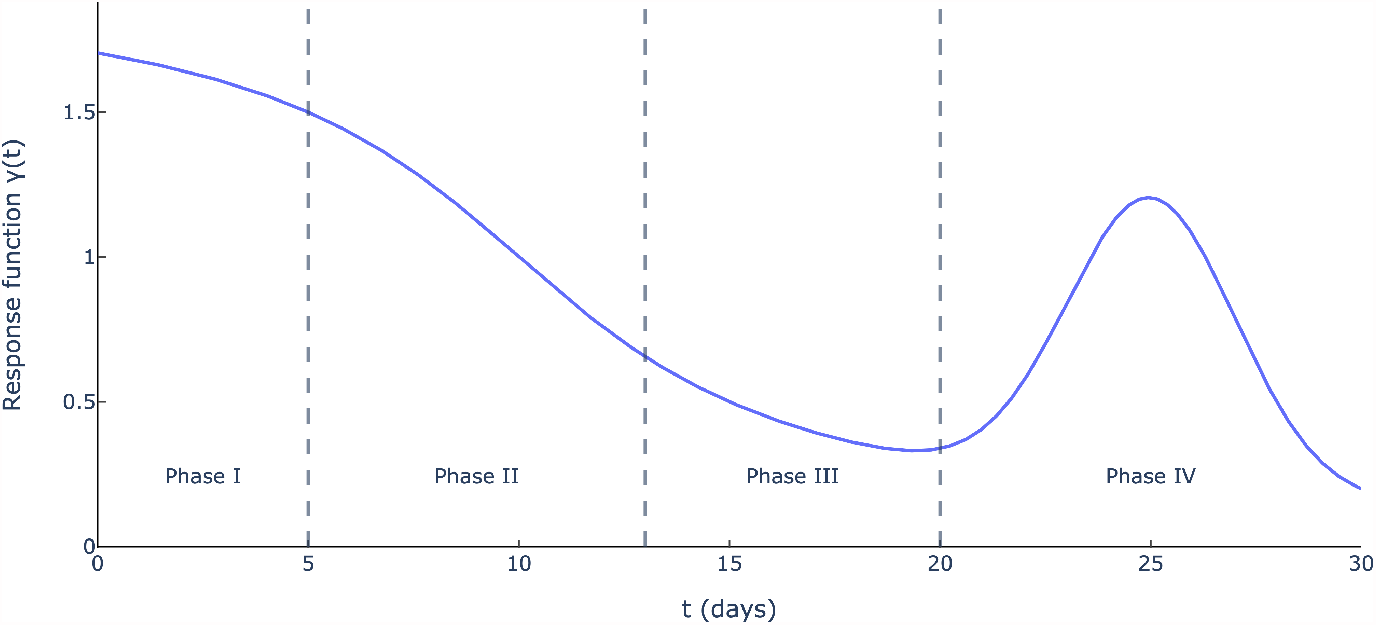
Illustration of 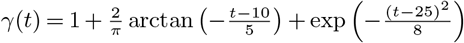 (i.e., 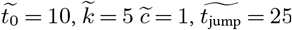, and 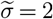).

Parameters 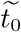 and 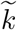 control the timing of such measures and the rapidity of their penetration, while the 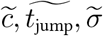 control the timing, magnitude, and duration of the resurgence. This formulation allows us to model, under the same framework, a wide variety of policies that different governments impose, including social distancing, stay-at-home policies, quarantines, and others, along with the societal response. This modeling captures the increasing force of intervention in the early-mid stages of the epidemic, and also accounts for the relaxation in behavior and measures in the later stages of the epidemic.

## 3. Results and Performance Analysis

In this section, we present the results of the DELPHI predictive model and its performances in terms of Mean Absolute Percentage Error (MAPE) and Root Mean Squared Error (RMSE) across time and regions, and benchmark it against the state-of-the-art COVID-19 models used by the CDC. We also analyze the sensitivity of DELPHI to perturbations in its parameters.

### 3.1. Forecasting Results

DELPHI was created in early April and has been continuously updated to reflect new observed data. The codebase is available on GitHub^1^ with the primary model being written in Python 3.7 using the SciPy and NumPy libraries. The implementation is also multiprocessing-friendly, which allows it to scale easily (using servers/machines with multiple CPUs/threads) to the high number of areas the model is fitted on every day. Figures 3a and 3b show our projections of the number of cases in Russia and the United Kingdom made on three different dates, and compare them against historical observations. These countries were chosen for illustrative purposes, as they are major countries with very different curves for the cases on the three evaluation dates. The results for Russia and the UK are however consistent with the overall performances across all countries of all regions, as described extensively in the rest of the section. They suggest that DELPHI achieves strong predictive performance, as the model has been consistently predicting, with high accuracy, the overall spread of the disease for several weeks. Notably, DELPHI was able to anticipate, as early as April 17th, the dynamics of the pandemic in the United Kingdom (resp. Russia) up to May 12th. At a time when 100-110K (resp. 30-35K) cases were reported, the model was predicting 220-230K (resp. 225-235K) cases by May 12th—a prediction that became accurate a month later. In the case of Russia, DELPHI was able to predict that the country was going to become a global hotspot as well as to accurately estimate the magnitude of the first wave of the outbreak even at an early stage of the pandemic (less than 0.025% of the population infected, vs more than 0.16% a month later which has put the country at the 4^*th*^ rank worldwide in terms of cumulative number of cases).

**Figure 3.**
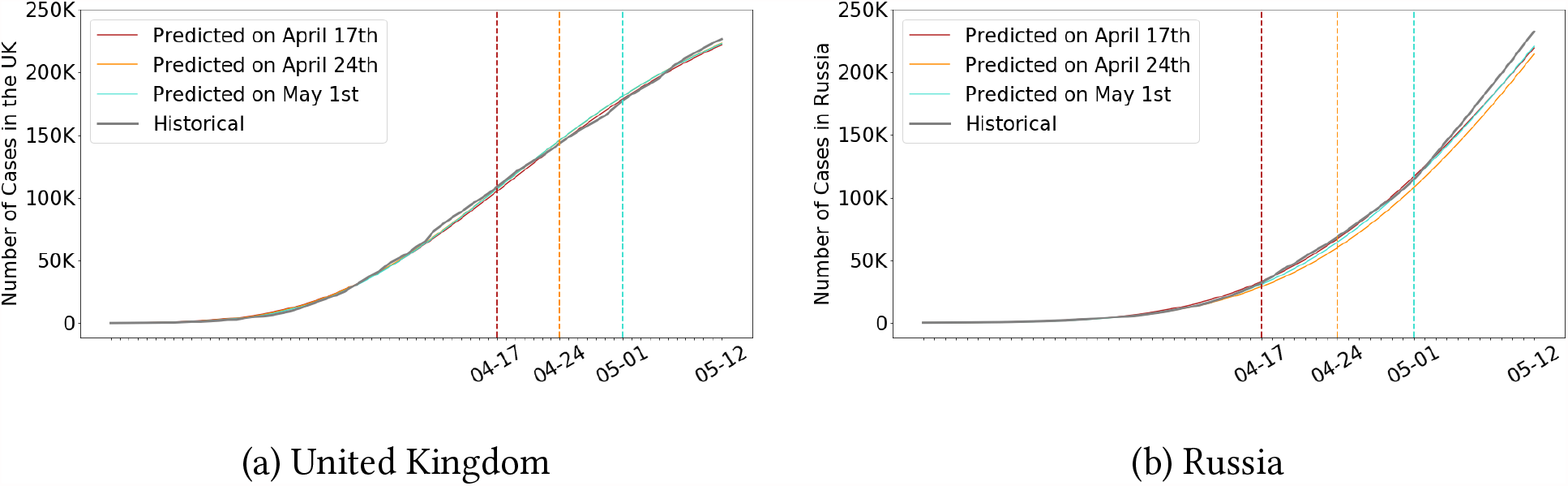
Cumulative number of cases in the UK (a) and Russia (b) according to our projections made at different points in time, against actual observations. Note there predicted curves largely overlap with the actual curve.

Furthermore, Table 2 reports the median Mean Absolute Percentage Error (MAPE) and the Root-Mean-Squared Error (RMSE) on the observed total cases and deaths in each area of the world for two periods: 1) The first period uses data up until April 27th, and evaluates on the 15 day period up until May 12th; 2) The second period uses data up to September 21st, and evaluates on the 15 day period up until October 6th. We illustrate the results over two separate periods to demonstrate the effect of the update in modeling to account for resurgence in infections and decay in the mortality percentage. Overall, our model seems to predict the epidemic progression relatively well in most countries across the two periods with *<* 10% MAPE on reported cases, and *<* 15% MAPE on reported deaths with a very competitive worldwide median MAPE at 5.8% for cases and 10.6% for deaths. Additionally, the areas with the highest MAPE are often those that have the fewest deaths, with selected examples in Table 1. This stems from the fact that DELPHI—like all SEIR-based models—is not designed to perform well on areas with small populations and interactions. The effect is further exacerbated by the choice of the metric, since MAPE inherently heavily penalizes errors on small numbers. If we turn our attention to RMSE, we see that the median RMSE in deaths in both periods were *<* 100 across all regions, which is significant given the high amount of deaths reported in each region (e.g. by the second period a majority regions in North America and Europe were reporting over 5, 000 cumulative deaths). The median RMSE on cases are more significant at just above 2, 000 cases in the second period, but is still small compared to the daily median number of recorded cases across all areas (∼ 60, 000).

**Table 1.**
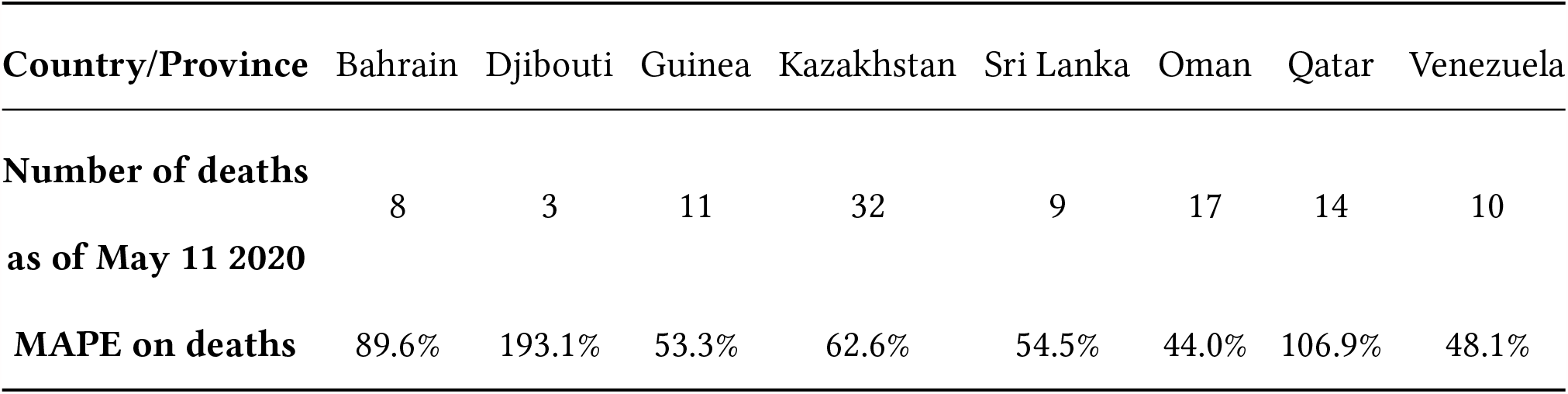
Breakdown of number of deaths vs. MAPE on deaths for large errors on the prediction period of April 28th to May12th.

| Country/Province | Bahrain | Djibouti | Guinea | Kazakhstan | Sri Lanka | Oman | Qatar | Venezuela |
| --- | --- | --- | --- | --- | --- | --- | --- | --- |
| **Number of deaths as of May 11 2020** | 8 | 3 | 11 | 32 | 9 | 17 | 14 | 10 |
| **MAPE on deaths** | 89.6% | 193.1% | 53.3% | 62.6% | 54.5% | 44.0% | 106.9% | 48.1% |

**Table 2.** Median country-level Mean Absolute Percentage Error (MAPE) and Root Mean Squared Error (RMSE) of the predicted number of cases and deaths in each region. Projections in the first (resp. second) half of the table are made using data up to 04/27 (resp. 09/21) for the period from 04/28 to 05/12 (resp. 09/22 to 10/06).

|  |  | Median MAPE Cases | Median MAPE Deaths | Median RMSE Cases | Median RMSE Deaths |
| --- | --- | --- | --- | --- | --- |
| Region | # Areas | (10th, 90th percentile) | (10th, 90th percentile) | (10th, 90th percentile) | (10th, 90th percentile) |
| April 28th |  |  |  |  |  |
| Africa | 19 | 14.7% (3.1, 32.0) | 23.4% (11.8, 60.3) | 138.7 (27.0, 1019.6) | 4.8 (1.1, 27.5) |
| Asia | 32 | 4.8% (2.1, 18.4) | 14.4% (2.9, 65.2) | 677.3 (51.5, 9778.1) | 13.0 (1.0, 151.6) |
| Europe | 42 | 3.4% (0.8, 12.9) | 9.0% (2.3, 24.3) | 238.1 (15.4, 3276.4) | 14.8 (1.6, 236.2) |
| North America | 10 | 7.9% (3.9, 28.3) | 12.6% (2.8, 23.6) | 594.4 (36.1, 1947.5) | 16.3 (4.0, 132.3) |
| Oceania | 2 | 3.2% (2.4, 4.1) | 12.0% (11.0, 13.0) | 68.6 (49.4, 87.8) | 2.0 (1.8, 2.3) |
| South America | 11 | 14.9% (7.6, 26.7) | 6.1% (3.3, 30.1) | 683.6 (31.4, 10815.0) | 6.8 (0.6, 426.1) |
| United States | 51 | 8.5% (1.9, 16.7) | 7.8% (3.3, 25.1) | 1231.6 (73.8, 4861.8) | 33.5 (1.7, 210.7) |
| World | 167 | 5.8% (1.5, 22.6) | 10.6% (2.9, 36.6) | 412.6 (26.1, 4788.7) | 12.0 (1.3, 193.1) |
| September 22nd |  |  |  |  |  |
| Africa | 53 | 5.2% (0.6, 30.4) | 4.2% (0.0, 42.6) | 364.5 (27.5, 2913.2) | 8.8 (0.0, 121.7) |
| Asia | 38 | 6.5% (1.7, 38.4) | 8.3% (1.4, 26.8) | 6311.3 (120.9, 53046.7) | 47.4 (0.7, 719.5) |
| Europe | 44 | 13.4% (3.8, 38.5) | 7.7% (1.1, 22.5) | 4452.9 (326.6, 22405.9) | 43.6 (1.7, 776.5) |
| North America | 14 | 7.3% (1.2, 17.1) | 4.8% (1.0, 22.0) | 2180.4 (187.5, 8656.7) | 48.9 (2.0, 269.0) |
| Oceania | 3 | 2.7% (1.6, 4.2) | 1.1% (0.8, 7.1) | 51.0 (16.0, 127.5) | 0.8 (0.6, 3.8) |
| South America | 13 | 9.4% (1.2, 15.5) | 5.2% (2.1, 22.7) | 5683.8 (473.2, 131184.4) | 95.6 (3.5, 4438.1) |
| US | 51 | 5.2% (1.3, 20.6) | 3.0% (0.5, 15.0) | 5323.5 (661.6, 17571.2) | 66.9 (3.1, 275.0) |
| World | 216 | 6.5% (1.2, 28.2) | 4.8% (0.6, 27.6) | 2170.0 (69.3, 18549.4) | 31.2 (1.1, 505.3) |

We further observe that after introducing additional terms to account for the decay in mortality percentage, the MAPE for deaths dropped significantly to 4.8% over the second period, demonstrating the efficacy of improved modeling as additional data became available.

### 3.2. Comparison with Other Models

To further understand and showcase the performance of DELPHI, we compare DELPHI to other top-performing models submitted to the CDC ensemble forecast in the context of predicting the number of deaths in the United States 4 weeks ahead, the furthest away and most difficult endpoint that the CDC ensemble forecast records. In particular, we have selected comparison models that are consistently included the CDC ensemble forecasts. This includes the models from University of Texas, Austin (UT-Mobility, **?**), Institute for Health Metrics and Evaluation (IHME-CurveFit, IHME (19)), Youyang Gu (YYG-ParamSearch, **?**), Northeastern University’s Laboratory for the Modeling of Biological and Sociotechnical Systems (MOBS-GLEAM COVID, **?**), Predictive Science Inc. (PSI-DRAFT, DRA (2020)), Los Alamos National Laboratory (LANL-GrowthRate, LAN (2020)), and Notre Dame University (NotreDame-mobility, Perkins and Espana (2020)).

In Figure 4a, we compare the out-of-sample MAPE of these models for the endpoint defined above using the actual weekly predictions submitted to the CDC between July and September. This particular period was selected because it encompassed the majority of the period of resurgence in the United States and its decay, making its prediction even more difficult. We observe that DELPHI consistently achieves low MAPE, and that its predictions are stable with a MAPE never exceeding 3.5% throughout the entire period. Figure 4b further illustrates the performance of DELPHI in comparison to other models by graphing the weekly ranking (with respect to MAPE). We observe that DELPHI consistently outperforms all other models, holds the first rank for 6 out of 13 weeks, and never drops below rank 4 among the 8 models evaluated: this provides further evidence of the real-world performance of DELPHI.

**Figure 4.**
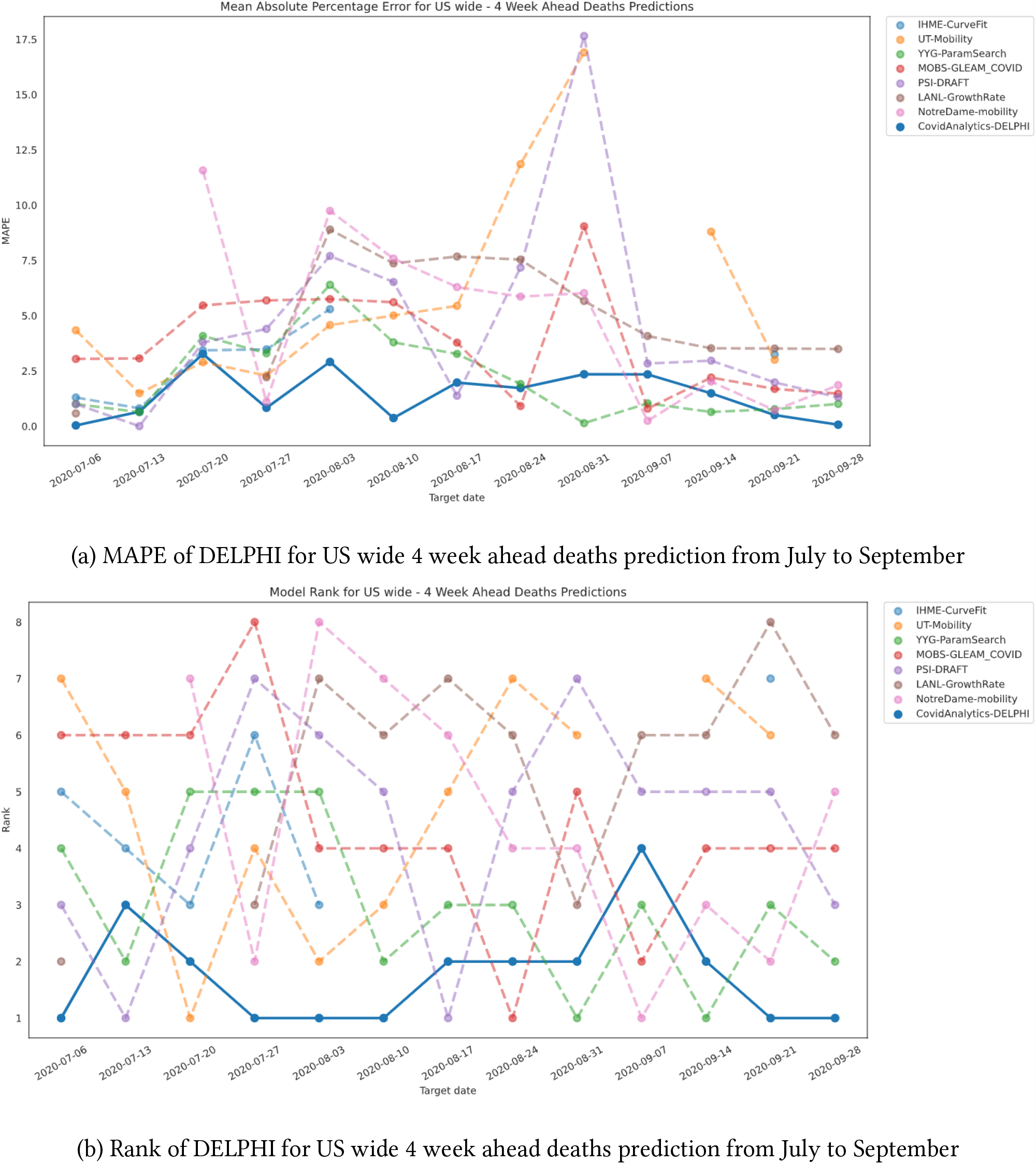
Comparison of 4 week MAPE on deaths prediction in the US between DELPHI and other models used by the CDC

### 3.3. Sensitivity Analysis

The DELPHI model utilizes a number of fixed parameters derived from literature or our clinical database in an attempt to reduce the amount of data needed for fitting the model. In reality, these parameters would be changing over time and therefore it is critical to understand the effect of fixing these parameters on the model. Therefore, we conducted an extensive sensitivity analysis to illustrate the effect on the model when the key fixed parameters are perturbed. Specifically, for every fixed parameter in DELPHI (*β, r*_*d*_, *σ, κ, p*_*d*_, *p*_*h*_), we randomly perturb the parameter by a normal noise term *ε* that has mean 0 and standard deviation of 20% of the nominal parameter’s absolute value (*i*.*e. ε* ∼ 𝒩 (0, 0.2 · |param|)). Then we fit the DELPHI model using data up to a certain prediction date using the modified fixed parameter and record the 30 day out-of-sample MAPE and compare with the MAPE of the actual model that was ran on that prediction date (using the publicly available records of our predictions).

We conducted the sensitivity experiment for all states in the United States and 6 international countries with large outbreaks: Italy, Spain, Brazil, South Africa, Japan and Russia. The countries were chosen to have a broad representation around the world. For completeness purposes, for each area we further conduct the experiment on 3 separate prediction dates. Figure 5a and 5b record the quantile plots of the absolute difference between the MAPE of the actual model and the perturbed model across all 6 perturbed parameters and 3 prediction dates (meaning that each box contains the distribution of 57 points for a given prediction date and perturbed parameter). We observe that for all 6 parameters, across both cases and deaths, the effect of the perturbation on the one-month MAPE is relatively small, with interquartile range mostly falling between ±5% for a perturbation with a standard deviation of 20% of the parameter value. This demonstrates that the results from the DELPHI model are robust to a (relatively important) change to the underlying fixed parameters, which comforts us in some of our core hypotheses.

**Figure 5.**
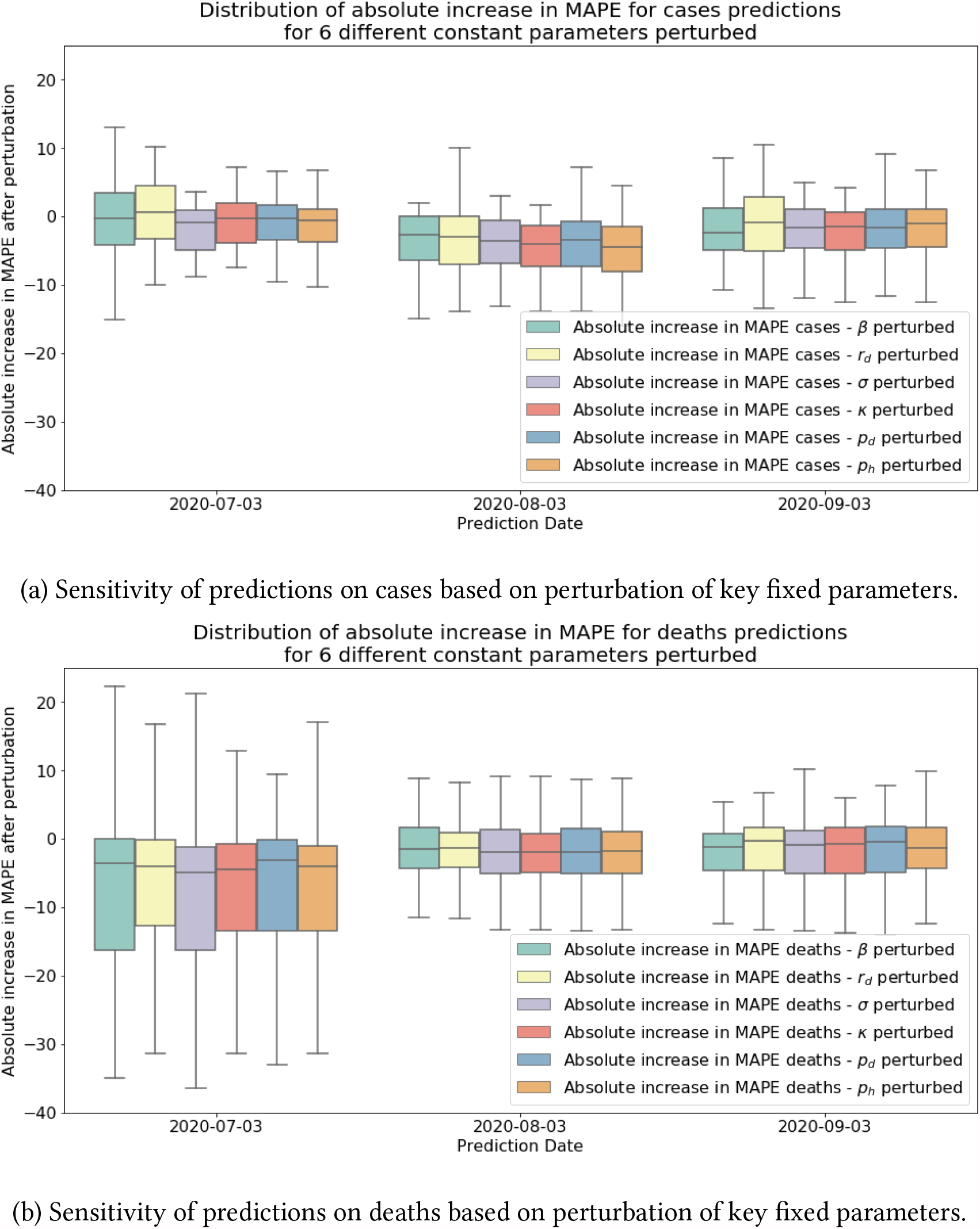
Sensitivity analysis of various fixed parameters, comparing perturbed MAPE on cases and deaths to their nominal counterparts without perturbations.

## 4. Evaluating Different Government Intervention Scenarios

In this section, we extend DELPHI to evaluate the impact of government interventions. That allows us to quantify the efficiency of Non-pharmaceutical Interventions (NPIs) and predict counterfactuals and “what-if” scenarios under different policies, which enable policy-makers to assess their COVID-19 response and decide on their future interventions accordingly.

### 4.1. Effect of Government Interventions

A particular benefit of the parametric modeling utilized by DELPHI is that it could easily be utilized for policy evaluation. For that, we can extract the normalized fitted government response curve *γ*(*t*) in each area, and utilize it to understand the impact of specific government policies that have been implemented. In particular, we aim to understand the average effect of each policy on *γ*(*t*) during the early stages of the pandemic to retrospectively understand which policies were effective. To this end, for all countries except the US, we collect data from the Oxford Coronavirus Government Response Tracker for historical data on government policies Hale et al. (2020), during the period between January 1st 2020, and May 19th 2020. For the US, we collect the policy data from the Institute for Health Metrics and Evaluation Murray et al. (2020) during the same period.

At each point in time, we categorize the government intervention data based on whether they restrict mass gatherings, schools, travel and work activities. We group travel restrictions and work restrictions together due to their tendency to be implemented simultaneously. From January 1st to May 19th, the 167 areas in total implemented 5 combinations of such interventions. Specifically, these are: (1) *No measure*; (2) *Restrict travel and work only*; (3) *Restrict mass gatherings, travel and work*; (4) *Restrict mass gatherings, schools, travel and work*; and (5) *Stay-at-Home*. The detailed correspondence between raw policy data and our categories are contained in the Appendix. Other potentially feasible combinations were not implemented by the countries, meaning that these 5 policies are mutually exclusive and collectively exhaustive. Then for each policy category *i* = 1, …, 5, we extract the average value of *γ*(*t*), 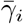, across all time periods and areas for which policy *i* was implemented. Then we calculate the residual fraction of infection rate under policy *i, p*_*i*_, compared to the baseline policy of no measure:

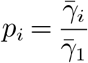

Table 3 shows the number of area-days that each policy was implemented around the world and its effect. We further report the standard deviation of such estimate treating each geographical area as an independent sample. We see that each selected policy was enacted for at least hundreds of Area-Days worldwide, while the stringent stay-at-home policy was cumulatively implemented the most. In particular, we see that mass gathering restrictions generate a large reduction in infection rate, with the incremental reduction between travel and work restrictions compared to mass gathering, travel, and work restrictions is 29.9 ± 6.9%. This is further supported by the large residual infection rate of 88.9 ± 4.5% when travel and work restrictions are implemented, but mass gatherings are allowed. Additionally, we observe that closing schools also generate a large reduction in the infection rate, with an incremental effect of 17.3 ± 6.6% on top of mass gathering and other restrictions. Stay-at-home orders produced the strongest reduction in infection rate across the different countries, with a residual infection rate of just 25.6 ± 3.7% compared to when no measure was implemented.

**Table 3.**
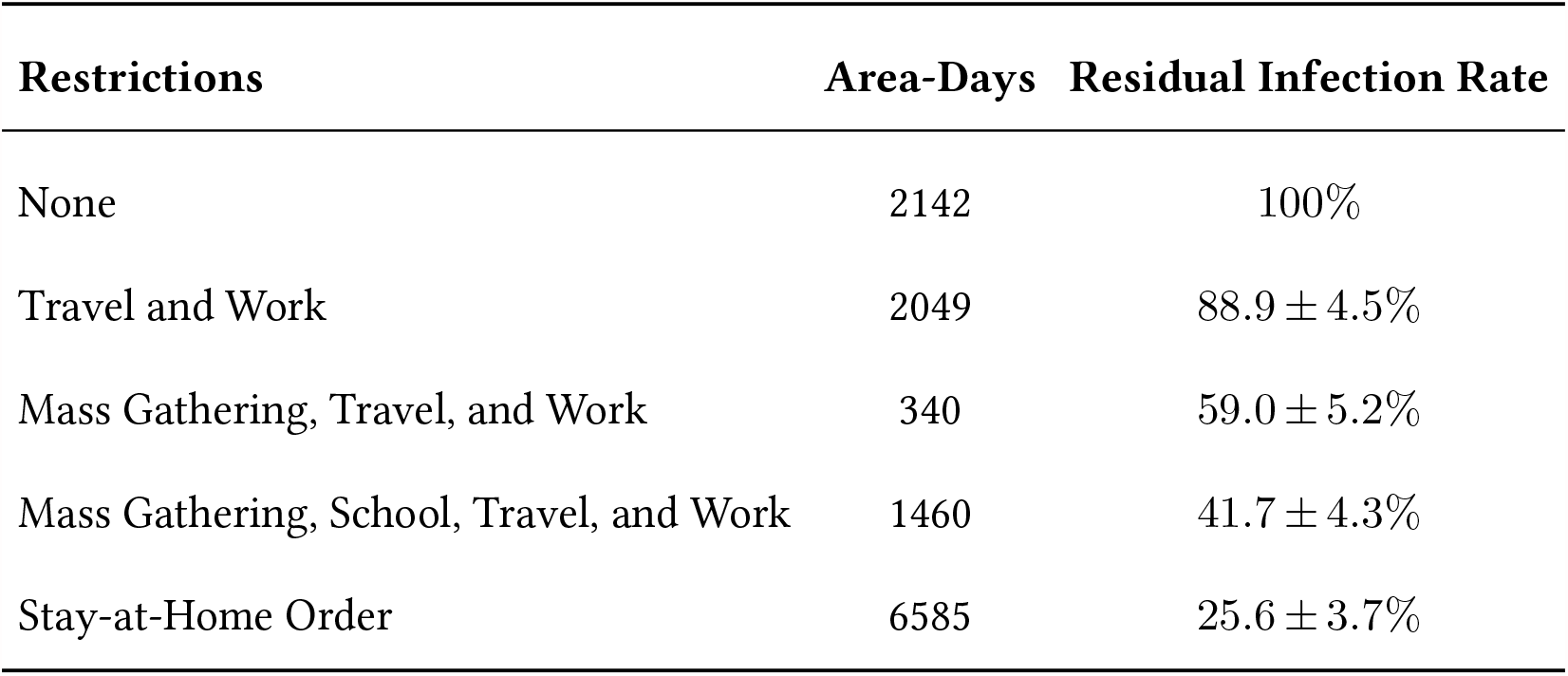
Implementation Length and Effect of each policy category as implemented across the world.

If COVID-19 has an average basic reproductive number *R*_0_ of 2.5-3 (Zhang et al. (2020), Liu et al. (2020a)), then on average, only the strongest measure (Stay-at-Home orders) are sufficient to control a COVID-19 epidemic in reducing *R*_0_ to be less than 1. However, this presents a serious dilemma to policymakers as the stay-at-home orders in early 2020 proved to carry a large social and humanitarian cost. This demonstrates that in the face of an already fast-growing outbreak, policymakers are often stuck between unpalatable policy options: in this case whether to let the pandemic grow uncontrollably or to inflict a significant humanitarian cost on society. In particular, this further stresses the value of early control for an epidemic as demonstrated in Section 4.2.

### 4.2. Modeling Alternative Initial Responses

Beyond analyzing the effect of different governmental interventions, our flexible parametric formulation allows us to easily explore alternative scenarios, such as what would have happened if the initial government interventions were enacted at different times. Specifically, to model what would happen if the government response was enacted *m* days earlier, we can simply correct the government response as followed:

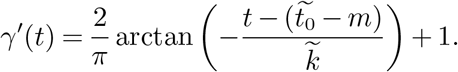

Figure 6 shows the percentage of cases and deaths avoided around the world by May 17th if the government interventions occurred one week earlier. The 50 countries with the highest reduction in deaths and cases are selected for clarity. The May 17th endpoint is selected as the vast majority of countries had not reopened by then, and thus the DELPHI assumption of increasing government intervention still holds. We can see that the DELPHI model predicts an over 75% reduction in both cases and deaths for many countries around the world if the restrictions were just implemented one week earlier. In particular, Western European countries such as Switzerland, Spain, and Italy would have benefited the most if restrictions had been enacted earlier. This corroborates the fact that these locations had some of the first outbreaks outside of Asia, and thus did not have as much time to react as countries which suffered a later outbreak, such as Romania and Iceland. Cumulatively across the world, DELPHI predicts that over 280,000 deaths or 68% of the world’s cumulative death count, could have been avoided around the world by May 17th, if every country in the world enacted its restrictions one week earlier.

**Figure 6.**
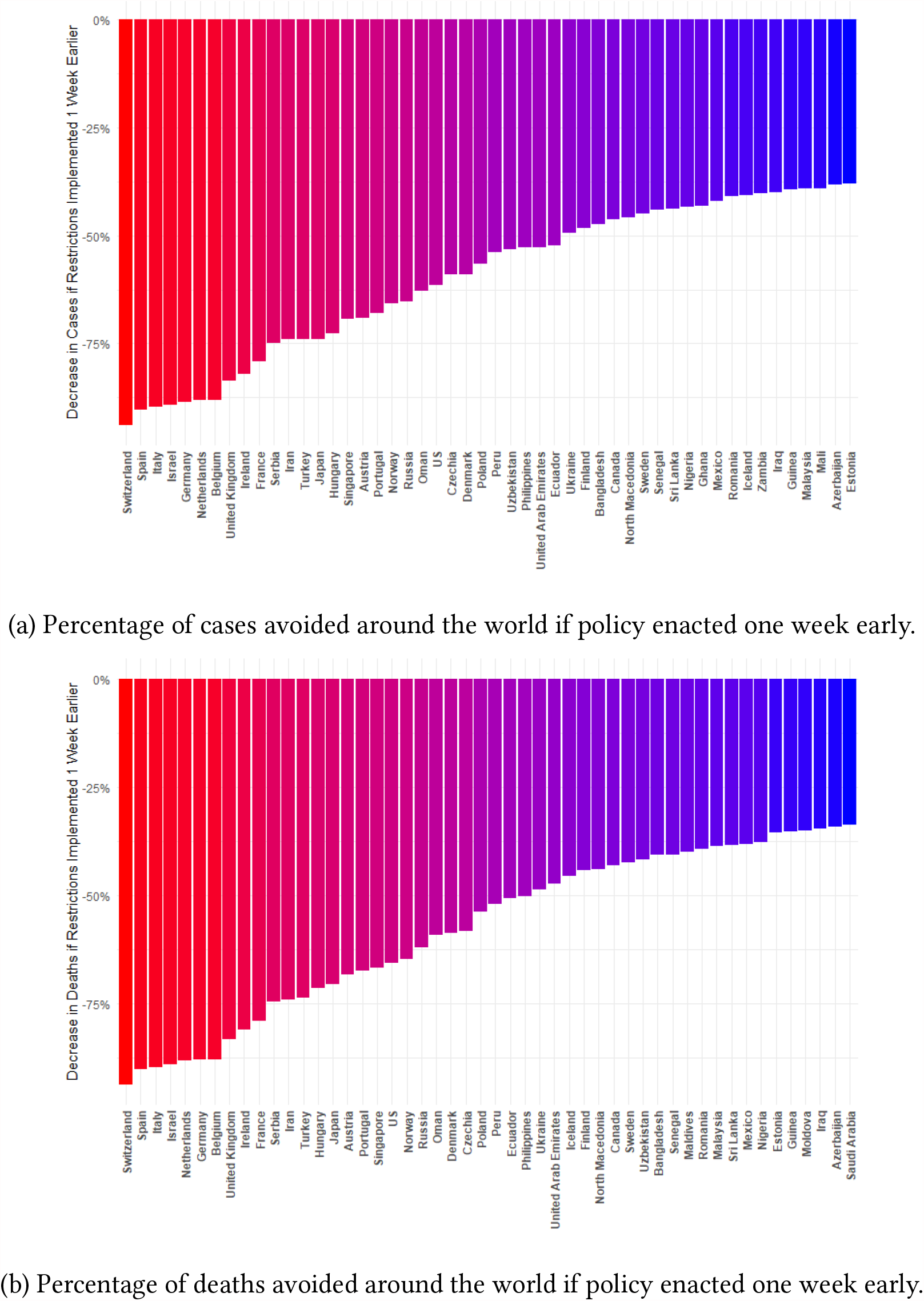
Scenario analysis if restrictions implemented one week earlier.

Another insightful scenario to consider is what would have happened if there had been no societal and governmental action against the epidemic, and COVID-19 was allowed to spread freely in the society. This can be naturally modeled in DELPHI with a constant government intervention curve of:

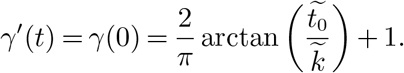

Using such response, DELPHI predicts that over 14.8 million individuals would have perished due to COVID-19 by May 17th if there had been a total lack of intervention. This demonstrates that the severe measures implemented by governments and societies worldwide saved a significant portion of the population around the world.

## 5. Analysis of What-If Scenarios for Long-term Planning

In Section 4.2, we observed how DELPHI could evaluate the effect of alternative initial responses by the governments worldwide. By extension, a natural use case of the DELPHI model is to create what-if scenarios due to changing *future* restrictions in different countries to enable long-term planning. In particular, in this section, we would illustrate how DELPHI was utilized by Janssen Pharmaceuticals in late May to create what-if scenarios on the effect of lifting restrictions in different countries in order to plan their Phase III trial location for their leading vaccine candidate, Ad26.Cov-2.S. By reverting the effect of each policy on *γ*(*t*) at the time of the hypothetical policy relaxation, Janssen Pharmaceuticals is able to understand the potential differing infection scenarios so that it can choose trial locations that would maximize the infection incidence in the placebo group. Since its successful deployment in late May, DELPHI has been continually utilized in further COVID-19 vaccine trials in the Janssen portfolio.

Specifically, suppose that we are considering a policy easing from policy *i* to *j* at time *t*_*c*_ in some area that has not yet experienced a resurgence 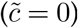. Then for all times *t* ≥ *t*_*c*_, we correct the government response as follows:

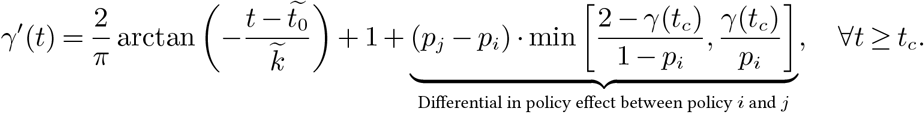

Essentially, we apply a correction term that is proportional to the fractional difference in policy effect between policy *i* and *j* (which is *p*_*j*_ − *p*_*i*_ *>* 0 as it is an easing). The multiplicative factor min 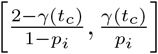 scales the fractional difference so that the resulting *γ′*(*t*_*c*_) is constrained within the initial range [0, 2]. Then, we would replace *γ*(*t*) with *γ′* (*t*) in the DELPHI model to forecast the epidemic under the updated policy. Using this correction factor, we predict what would happen in different areas under various future policies. Figure 7 shows results for France and Brazil respectively, under policy change implemented on June 16th (four weeks after the last historical value on May 19th). Further results for other countries are contained in the Appendix.

**Figure 7.**
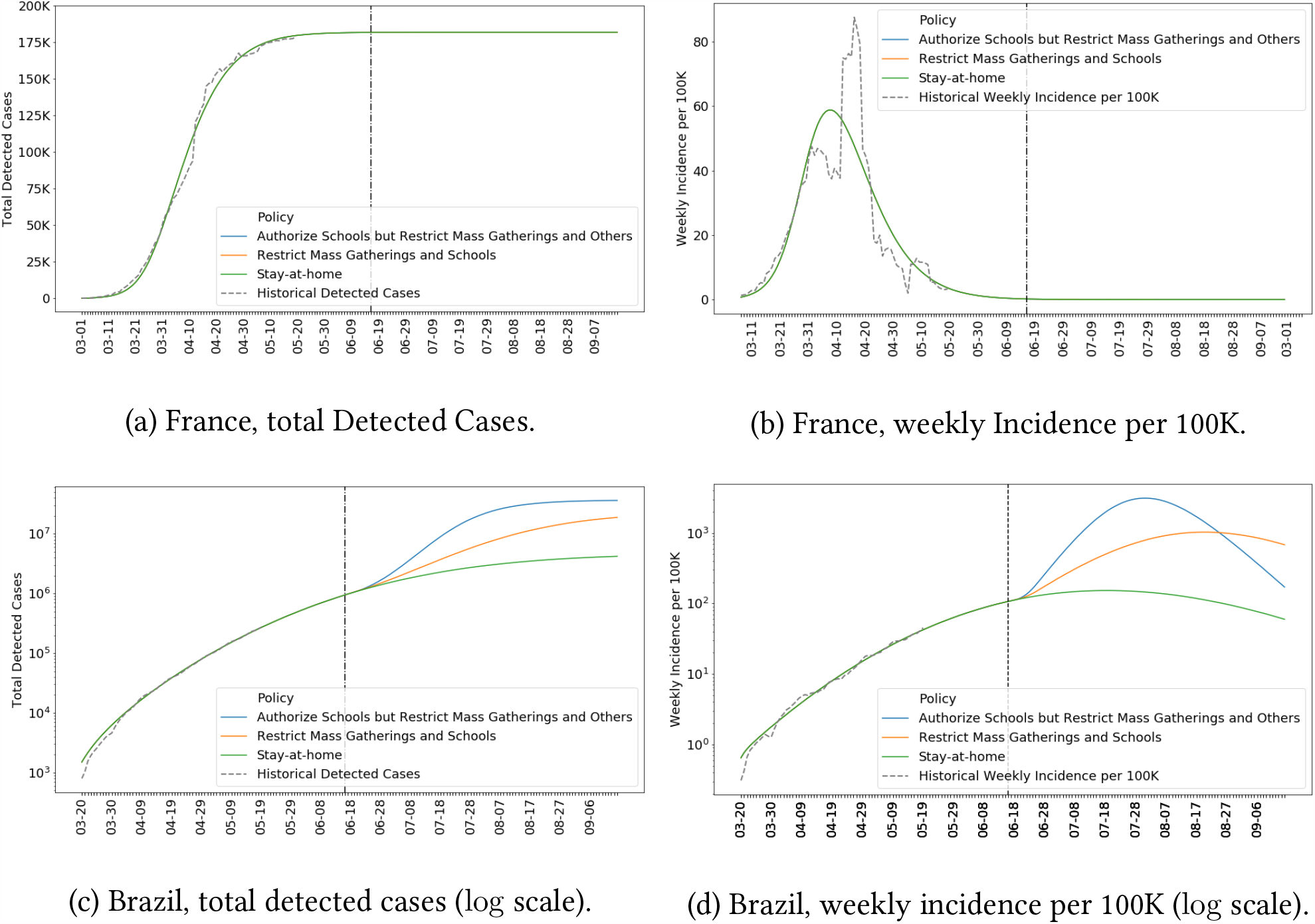
Forecasts of total detected cases and weekly incidence per 100K for France and Brazil under various policies.

We observe different levels of risk for the same re-opening strategies across different countries. For example, Figure 7c predicts that loosening measures in Brazil on June 16th would result in a second wave of infections with up to 6.8 million additional cases by July 15th, while even a stay-at-home order would lead to almost 1.9 million additional cases. Such alarming numbers can be understood through Figure 7d where we compute a rolling average of the weekly incidence of cases per 100K people. We can see that Brazil is still on a steep ascending curve, and that any kind of loosening could be catastrophic. Such behaviour stands in sharp contrast with France’s situation. Figure 7b demonstrates that the peak has long passed in France and the epidemic has mostly died out. Thus, as we can see in Figure 7a, loosening policies (like France has already started doing) is likely to only minimally affect the number of infections.

To further understand the disparate impact of the policies across countries, we made predictions for the situation around the world assuming a policy that involves mass gathering, travel, and work restrictions was universally implemented on 06/16. Figure 8a shows three clusters of countries for July 15th:

**Figure 8.**
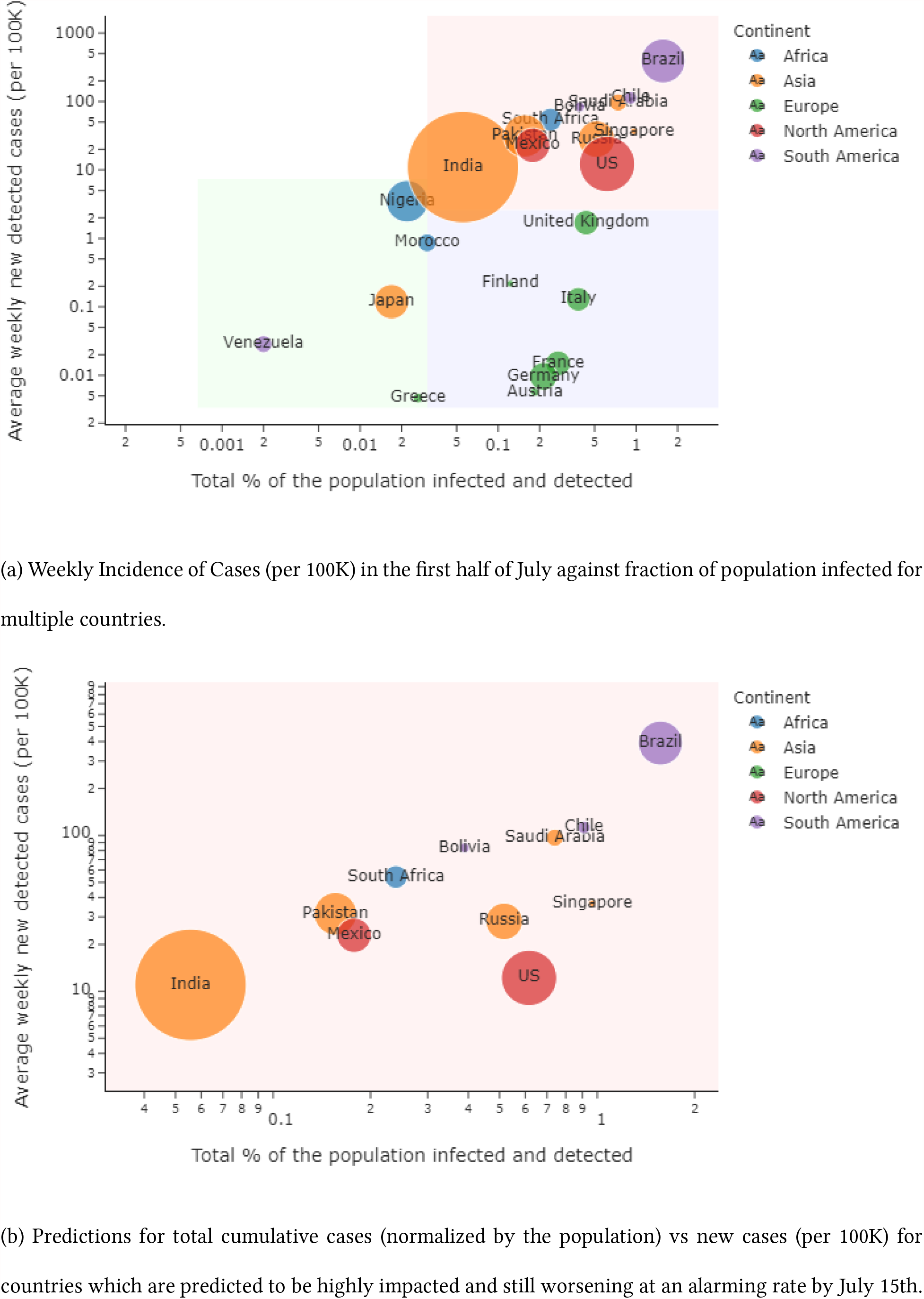
World Predictions for Early July under Mass Gathering, Travel and Work Restrictions.

- Countries with a small number of cumulative cases (relative to the population), and that are in a late stage of the pandemic with relatively few new cases, such as Greece, Japan, Morocco and Venezuela.
- Countries with a large number of cumulative cases, but that are in a late stage of the pandemic, with relatively few new cases, mainly in Western and Northern Europe (e.g. the United Kingdom, Italy, France and Finland).
- Countries where the pandemic has had a large impact with a large number of cumulative cases, and where the situation will still be worsening at an alarming rate. These include the United States, India and Brazil. A close-up of these countries is presented in Figure 8b, where we see that DELPHI predicts Brazil would be severely hit by July, with up to 8% of the entire population infected, if the hypothetical policy above is implemented. This suggests that in these countries, such hypothetical policy could be inadequate for controlling the epidemic, and a stronger policy (such as Stay-at-Home orders) is needed.

## 6. Limitations

One limitation of this analysis is that some parameters are fixed to a constant value, most importantly the median time to leave incubation *T*_*β*_ (fixed at 5 days), the median time to recovery (*T*_*σ*_ fixed at 10 days for non-hospitalized patients and *T*_*κ*_ fixed at 15 days for hospitalized patients) and the detection percentage fixed at 20%. However, the detection times are reasonably consistent throughout a certain number of studies (Lauer et al. (2020), Hu et al. (2020), Kluytmans et al. (2020), Liu et al. (2020b), Grein et al. (2020)). For the detection rate, using the sensitivity analysis and the values inferred from random serology testing, we can show that the DELPHI model and its predictions are robust to this input. DELPHI also does not currently capture a potential time-varying nature of these fixed constants. Including such effect could sharpen the analysis further, though at the expense of increased fitting difficulty and data requirements. Furthermore, while we decided to use a decaying mortality percentage (using the arctan function) which allowed us to predict even more precisely deaths all around the world, one criticism could be that this parametric function cannot grasp a bump in mortality percentage. Indeed, in December 2020, the observed mortality percentage has started raising again in some regions of the world, mostly because of the temporary saturation of hospital systems: one way to account for this could be to use, similarly to what we did in the government intervention modeling, an exponential bump. While we haven’t seen the need to do so now, we acknowledge that this could be a limitation for a few limited periods.

One further important limitation of the DELPHI model is that it does not explicitly model the effect of asymptotic undetected infections, who are unlikely to quarantine and thus would participate in the infection loop throughout their entire infectious period (currently we assume that everyone participates in the infection loop for some time before they are quarantined in some fashion). This limitation is the result of the long-standing significant debate in the medical community regarding the magnitude of COVID-19 infections due to asymptomatic undetected individuals. There were early estimates (see e.g. Ing et al. (2020)) that suggest a high percentage of infections were caused by such effect, but a recent published meta-analysis (Byambasuren et al. 2020) concluded that asymptotic infections account for no more than 17% of total infections and asymptotic infections are 42% less likely to further infect other individuals, so the impact of such infections on the pandemic is small. Given such wide-ranging differing estimates of the effect, we made a decision not to explicitly model asymptotic undetected patients as there does not seem to be sufficient reliable data on their impact in the COVID-19 pandemic. However, once reliable data is present, an explicit modeling of such effect could further improve the forecasting results for DELPHI. It is also interesting to note that DELPHI has the same limitations than other SEIR-based models (Holmdahl and Buckee (2020)) in terms of size of populations and minimum number of cases. Experimentally, it translates in some rare cases with a significant drop in accuracy for very small provinces (as described in Table 1). Holmdahl and Buckee (2020) also point out that SEIR-based models, including DELPHI, struggle with long-term predictions: even though DELPHI is able to detect new waves very well when the occur, it absolutely cannot detect them beforehand, causing potentially the long-term predictions to be off if one or multiple new waves happen between the training and the prediction dates.

This analysis also assumes, in analyzing government interventions, that the same nominal policy (e.g. Mass gathering restrictions) could be compared across countries. In reality, different countries have implemented variants (though largely similar) of restrictions under the same name, and this could further impact the validity of the analysis.

In the reopening analysis, we have assumed that the effect of government interventions imposed at the start of the epidemic is indicative of the effect when it is removed. This is potentially affected by a permanent change in social behavior during the epidemic. For example, if a significant portion of the population adapts social distancing measure even after the official restrictions are lifted, this could lead to a smaller resurgence of infections than what is predicted in the analysis.

## 7. Conclusions

We introduced DELPHI, a novel epidemiological model that extended SEIR to include many realistic effects critical in this pandemic. DELPHI was able to accurately predict the spread of COVID-19 in many countries, and aid planning for many organizations worldwide, among which governmental entities and pharmaceutical companies. Furthermore, the explicit modeling of government interventions allowed us to understand their effect, and help inform how societies could reopen.

## Data Availability

All data used in this paper is available at the DELPHI repository hosted on github.

https://github.com/CSSEGISandData/COVID-19

https://covid19.healthdata.org/united-states-of-america

https://www.bsg.ox.ac.uk/research/research-projects/coronavirus-government-response-tracker

https://github.com/COVIDAnalytics/DELPHI

## Appendices

### A. Detailed Forecasting Results for the World

**Figure A1.**
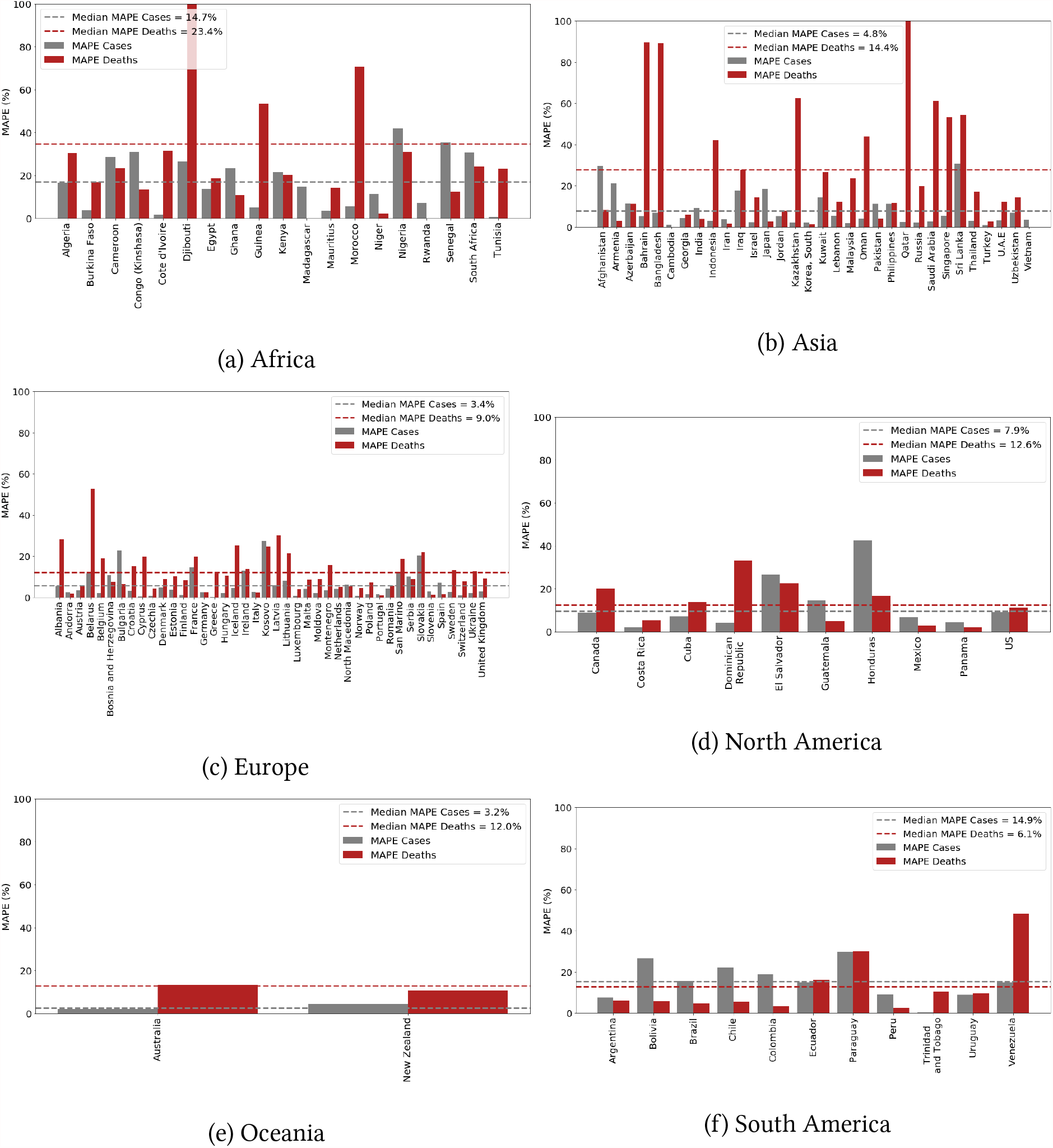
Mean Absolute Percentage Error (MAPE) of the predicted number of cases and deceases in each country (projections made using data up to 04/27 for the period from 04/28 to 05/12).

### B. Correspondence between Policy Data and Categorized Policies

**Table A1.** Correspondence between Oxford Policy Data and Categorized Policies.

| Oxford Policy | Categorized Policy |
| --- | --- |
| C3: Cancel Public Events<br>C4: Restrictions on gatherings<br>C5: Close public transport | Restrict Mass Gatherings |
| C2: Workplace closing<br>C7: Restrictions on internal movement<br>C8: International travel controls | Travel and Work Restrictions |
| C1: School closing | School Restrictions |
| C6: Stay at home requirements | Stay-at-Home Order |

**Table A2.** Correspondence between IHME Policy Data and Categorized Policies.

| IHME Policy | Categorized Policy |
| --- | --- |
| Mass Gathering Restrictions | Restrict Mass Gatherings |
| Business Closure<br>Non-essential Businesses Closed<br>Travel Severely Limited | Travel and Work Restrictions |
| Educational Facilities Closed | School Restrictions |
| Stay-at-Home Order | Stay-at-Home Order |

### C. Additional Results for Reopening Strategies

**Figure A2.**
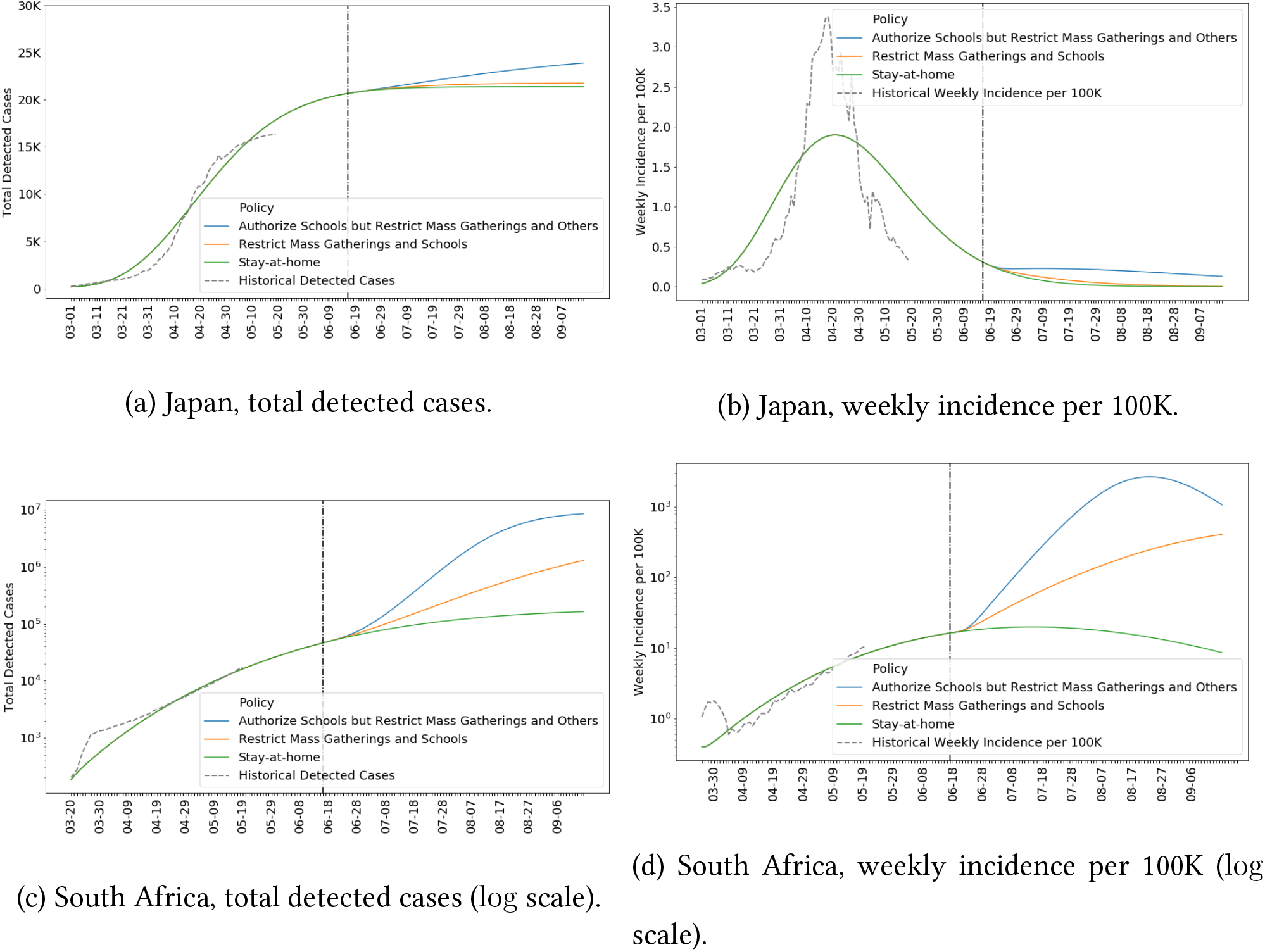
Forecasts of total detected cases and weekly incidence per 100K for Japan and South Africa under various policies.

## Notes

### Competing Interest Statement

The authors have declared no competing interest.

### Funding Statement

No Funding Sources.

### Author Declarations

No IRB approval required.

## References

(2020) Lanl covid-19 cases and deaths forecasts. URL https://covid-19.bsvgateway.org/.

(2020) Psi-draft. URL https://zoltardata.com/model/254.

Arons MM, Hatfield KM, Reddy SC, Kimball A, James A, Jacobs JR, Taylor J, Spicer K, Bardossy AC, Oakley LP, et al. (2020) Presymptomatic sars-cov-2 infections and transmission in a skilled nursing facility. New England Journal of Medicine.

Bendavid E, Mulaney B, Sood N, Shah S, Ling E, Bromley-Dulfano R, Lai C, Weissberg Z, Saavedra R, Tedrow J, et al. (2020) Covid-19 antibody seroprevalence in santa clara county, california. MedRxiv.

Bertsimas D, Bandi H, Boussioux L, Cory-Wright R, Delarue A, Digalakis V, Gilmour S, Graham J, Kim A, Lahlou Kitane D, Lin Z, Lukin G, Li M, Mingardi L, Na L, Orfanoudaki A, Papalexopoulos T, Paskov I, Pauphilet J, Skali Lami O, Sobiesk M, Stellato B, Carballo K, Wang Y, Wiberg H, Zeng C (2020) An aggregated dataset of clinical outcomes for covid-19 patients. URL http://www.covidanalytics.io/datasetdocumentation.

Breton T (2020) An estimate of unidentified and total us coronavirus cases by state on april 21, 2020. SSRN.

Byambasuren O, Cardona M, Bell K, Clark J, McLaws ML, Glasziou P (2020) Estimating the extent of true asymptomatic covid-19 and its potential for community transmission: systematic review and meta-analysis. Available at SSRN 3586675.

Chinazzi M, Davis JT, Ajelli M, Gioannini C, Litvinova M, Merler S, y Piontti AP, Mu K, Rossi L, Sun K, et al. (2020) The effect of travel restrictions on the spread of the 2019 novel coronavirus (covid-19) outbreak. Science 368(6489):395–400.

Dean NE, y Piontti AP, Madewell ZJ, Cummings DA, Hitchings MD, Joshi K, Kahn R, Vespignani A, Halloran ME, Longini Jr IM (2020) Ensemble forecast modeling for the design of covid-19 vaccine efficacy trials. Vaccine 38(46):7213–7216.

Doi A, Iwata K, Kuroda H, Hasuike T, Nasu S, Kanda A, Nagao T, Nishioka H, Tomii K, Morimoto T, et al. (2020) Estimation of seroprevalence of novel coronavirus disease (covid-19) using preserved serum at an outpatient setting in kobe, japan: A cross-sectional study. medRxiv.

Erikstrup C, Hother CE, Pedersen OBV, Mølbak K, Skov RL, Holm DK, Sækmose S, Nilsson AC, Brooks PT, Boldsen JK, et al. (2020) Estimation of sars-cov-2 infection fatality rate by real-time antibody screening of blood donors. medRxiv.

Grein J, Ohmagari N, Shin D, Diaz G, Asperges E, Castagna A, Feldt T, Green G, Green ML, Lescure FX, et al. (2020) Compassionate use of remdesivir for patients with severe covid-19. New England Journal of Medicine.

Hale T, Webster S, Petherick A, Phillips T, Kira B (2020) Oxford covid-19 government response tracker, blavatnik school of government. Oxford University. Creative Commons Attribution CC BY standard. Available at: https://www.bsg.ox.ac.uk/covidtracker. Accessed on: April 14:p2020.

Holmdahl I, Buckee C (2020) Wrong but useful — what covid-19 epidemiologic models can and cannot tell us. New England Journal of Medicine 383(4):303–305, URL http://dx.doi.org/10.1056/NEJMp2016822.

Hu Z, Song C, Xu C, Jin G, Chen Y, Xu X, Ma H, Chen W, Lin Y, Zheng Y, et al. (2020) Clinical characteristics of 24 asymptomatic infections with covid-19 screened among close contacts in nanjing, china. Science China Life Sciences 1–6.

IHME (19) health service utilization forecasting team, murray cjl. Forecasting COVID-19 impact on hospital bed-days, ICU-days, ventilator-days and deaths by US state in the next 4.

Ing AJ, Cocks C, Green JP (2020) Covid-19: in the footsteps of ernest shackleton. Thorax.

Kermack WO, McKendrick AG (1927) A contribution to the mathematical theory of epidemics. Proceedings of the royal society of london. Series A, Containing papers of a mathematical and physical character 115(772):700–721.

Kluytmans M, Buiting A, Pas S, Bentvelsen R, van den Bijllaardt W, van Oudheusden A, van Rijen M, Verweij J, Koopmans M, Kluytmans J (2020) Sars-cov-2 infection in 86 healthcare workers in two dutch hospitals in march 2020. medRxiv.

Krantz SG, Rao ASS (2020) Level of under-reporting including under-diagnosis before the first peak of covid-19 in various countries: Preliminary retrospective results based on wavelets and deterministic modeling. Infection Control & Hospital Epidemiology 1–8.

Lauer SA, Grantz KH, Bi Q, Jones FK, Zheng Q, Meredith HR, Azman AS, Reich NG, Lessler J (2020) The incubation period of coronavirus disease 2019 (covid-19) from publicly reported confirmed cases: estimation and application. Annals of internal medicine 172(9):577–582.

Liu Y, Gayle AA, Wilder-Smith A, Rocklöv J (2020a) The reproductive number of covid-19 is higher compared to sars coronavirus. Journal of travel medicine.

Liu Y, Sun W, Chen L, Wang Y, Zhang L, Yu L (2020b) Clinical characteristics and progression of 2019 novel coronavirus-infected patients concurrent acute respiratory distress syndrome. medRxiv.

Lourenço J, Paton R, Ghafari M, Kraemer M, Thompson C, Simmonds P, Klenerman P, Gupta S (2020) Fundamental principles of epidemic spread highlight the immediate need for large-scale serological surveys to assess the stage of the sars-cov-2 epidemic. medRxiv.

Mehrotra P, Ivan J (2020) Prophet logistic forecasting.

Murray C, et al. (2020) Forecasting the impact of the first wave of the covid-19 pandemic on hospital demand and deaths for the usa and european economic area countries. medRxiv.

Niehus R, Martinez de Salazar Munoz P, Taylor A, Lipsitch M (2020) Qyantifying bias of covid-19 prevalence and severity estimates in wuhan, china that depend on reported cases in international travelers. medRxiv.

Nocedal J, Wright S (2006) Numerical optimization (Springer Science & Business Media).

Peng L, Yang W, Zhang D, Zhuge C, Hong L (2020) Epidemic analysis of covid-19 in china by dynamical modeling. arXiv preprint arXiv:2002.06563.

Perkins A, Espana G (2020) Notredame-fred covid-19 forecasts.

Rodriguez A, Tabassum A, Cui J, Xie J, Ho J, Agarwal P, Adhikari B, Prakash BA (2020) Deepcovid: An operational deep learning-driven framework for explainable real-time covid-19 forecasting. medRxiv.

Sood N, Simon P, Ebner P, Eichner D, Reynolds J, Bendavid E, Bhattacharya J (2020) Seroprevalence of sars-cov-2– specific antibodies among adults in los angeles county, california, on april 10-11, 2020. JAMA.

Streeck H, Schulte B, Kuemmerer B, Richter E, Höller T, Fuhrmann C, Bartok E, Dolscheid R, Berger M, Wessendorf L, et al. (2020) Infection fatality rate of sars-cov-2 infection in a german community with a super-spreading event. medRxiv.

Wang C, Liu L, Hao X, Guo H, Wang Q, Huang J, He N, Yu H, Lin X, Pan A, et al. (2020) Evolving epidemiology and impact of non-pharmaceutical interventions on the outbreak of coronavirus disease 2019 in wuhan, china. medRxiv.

Wise J (2020) Covid-19: Surveys indicate low infection level in community.

Woody S, Tec MG, Dahan M, Gaither K, Lachmann M, Fox S, Meyers LA, Scott JG (2020) Projections for first-wave covid-19 deaths across the us using social-distancing measures derived from mobile phones. medRxiv.

Xiang Y, Sun D, Fan W, Gong X (1997) Generalized simulated annealing algorithm and its application to the thomson model. Physics Letters A 233(3):216–220.

Xu H, Huang S, Liu S, Deng J, Jiao B, Ai L, Xiao Y, Yan L, Li S (2020) Evaluation of the clinical characteristics of suspected or confirmed cases of covid-19 during home care with isolation: A new retrospective analysis based on o2o. Available at SSRN 3548746.

Zhang S, Diao M, Yu W, Pei L, Lin Z, Chen D (2020) Estimation of the reproductive number of novel coronavirus (covid-19) and the probable outbreak size on the diamond princess cruise ship: A data-driven analysis. International Journal of Infectious Diseases 93:201–204.

